# Spatial patterns of child mortality in Nanoro HDSS site, Burkina Faso

**DOI:** 10.1101/2020.07.06.20147645

**Authors:** Navideh Noori, Karim Derra, Innocent Valea, Assaf P. Oron, Aminata Welgo, Toussaint Rouamba, Palwende Romuald Boua, Athanase Some, Eli Rouamba, Edward Wenger, Hermann Sorgho, Halidou Tinto, Andre Lin Ouédraogo

## Abstract

**Background:** Half of global child deaths occur in sub-Saharan Africa. Understanding child mortality patterns and risk factors will help inform interventions to reduce this heavy toll. The Nanoro Health and Demographic Surveillance System (HDSS), Burkina Faso was described previously, but spatial patterns of child mortality in the district had not been studied. Similar studies in other districts indicated accessibility to health facilities as a risk factor, usually without distinction between facility types.

**Methods:** Using Nanoro HDSS data from 2009 to 2013, we estimated the association between under-5 mortality and accessibility to inpatient and outpatient health facilities, seasonality of death, and age group.

**Results:** Living in homes 40-60 minutes and >60 minutes travel time from an inpatient facility was associated with 1.52 (95% CI: 1.13-2.06) and 1.74 (1.27-2.40) greater hazard of under-5 mortality, respectively, than living in homes <20 minutes from an inpatient facility. No such association was found for outpatient facilities. Seasonality of death was significantly associated with under-5 mortality, and the wet season (July-November) was associated with 1.28 (1.07, 1.53) higher under-5 mortality than the dry season (December-June), likely reflecting the malaria season.

**Conclusions:** Our results emphasize the importance of geographical accessibility to health care, and also distinguish between inpatient and outpatient facilities.

## Background

Since the establishment of the Millennium Development Goals in 1990, there has been substantial progress in reducing child mortality globally, from 93 deaths in 1990 to 39 deaths in 2017 per 1000 live births. Nonetheless, an estimated 5.4 million children under age five died in 2017, out of which 2.5 million died during the first month of their life [1]. About half of child deaths occurred in sub-Saharan Africa [2]. In 2015, the Sustainable Development Goals (SDGs) were defined, aiming to reduce under-five mortality to below 25 per 1000 live births by 2030 [3]. To achieve these targets, urgent action in sub-Saharan Africa is needed, as well as higher-quality information to guide this action [4]. Among sub-Saharan countries, Burkina Faso, where our study area is situated, has made great progress in reducing under-5 mortality by about 58% from 201 to 84.6 deaths per 1,000 live births between 1990 and 2016, but this rate is still much higher than the SDGs [1].

To track progress towards child survival goals and to plan effective interventions for child health, identifying the major drivers of child mortality as well as data-driven estimates of child mortality are necessary [4]. However, countries with the highest child mortality burden lack civil registration and vital statistics (CRVS) systems accounting for all births, deaths and causes of death. In these countries, the location and timing of child deaths and the overall death rates, are highly uncertain. What we know about these crucial public-health questions is informed mostly by nationally representative surveys such as the Demographic and Health Surveys (DHS), conducted every several years.

A Health and Demographic Surveillance System (HDSS) is a local CRVS system that routinely monitors the health and demographic characteristics of a population living in a specific area. HDSS data facilitate detailed local studies of public health in general, and child mortality in particular. As of 2020, forty-nine HDSS sites participate in the International Network for the Demographic Evaluation of Populations and Their Health in Developing Countries (INDEPTH), recording the life events of over three million people in 17 African and Asian countries [5]. Several studies have investigated spatial [6, 7, 8, 9, 10], temporal [11, 12] and demographic [11, 13, 14] factors affecting child mortality in HDSSs. However, no study to date has analyzed such patterns in the relatively new Nanoro HDSS in rural north-central Burkina Faso.

Risk of child mortality varies over space and time, and it is important to identify the areas at the highest risk in order to focus intervention-based efforts in those areas. One source of spatial heterogeneity is accessibility to health facilities [15, 16]. Poor access to health care remains a concern in many low-income countries [17]. A growing number of studies have estimated the effect of distance from a health facility upon child mortality. The first meta-analysis of such studies was published in 2012 [16] and was updated more recently [18]. They found that living *>* 5 km away from a facility is associated with 62% higher neonatal mortality based on 4 studies, and 57% higher under-5 mortality based on 9 studies; both effects were deemed highly significant. In addition, a study aggregating 29 DHSs from 21 countries found that living *>* 10 km from a facility was strongly associated with 27% higher odds of neonatal mortality. Both the meta-analyses and the DHS-based study did not distinguish between smaller and larger facilities. Most above mentioned studies used simple Euclidean distance, or local expert opinion about distance or travel time, as the exposure variable. More sophisticated approaches to estimate real-life travel distance or time [22] have been published only rarely in this context.

Mortality also varies over time as a result of changes in health care-seeking, age and season of birth and death [11, 12], and environmental conditions [19]. In the Nouna HDSS, Burkina Faso, infants born during the rainy season were associated with higher mortality risk compared with those born during the dry season [11]. During the rainy season, flooded roads limit the access to health care, especially in the rural region. In most of West Africa, the rainy season also coincides with food shortage until the harvest arrives [11]. Seasonality also drives cause-specific mortality patterns due to malaria, pneumonia and diarrhea, which were the leading causes of child mortality in Burkina Faso in 2010 [20].

Here we present a quantitative investigation of local under-5 mortality patterns in the Nanoro HDSS site. We were particularly interested in identifying the drivers of spatial heterogeneity in child mortality risk in the area, in view of recent progress on global accessibility estimates capturing inequalities in infrastructural development.

## Methods

### Study Area and Data

Nanoro HDSS site was established in 2009 by the Clinical Research Unit of Nanoro (CRUN), located in the Centre Medicale Saint Camille de Nanoro (CMA), with the goal of evaluating population demography and health living conditions within the health district [24]. Nanoro is located about 85km from the capital city, Ouagadougou. The Nanoro Demographic Surveillance Area (DSA) lies within the health district of Nanoro and includes 24 villages. Initial census started from March to April 2009, and recorded housing and demographic characteristics of 54,781 individuals. Since then, census follow-up has been carried out every four months. Data collected at the individual level include births, deaths, pregnancies, in/out-migrations (temporary or permanent), and relationships (mother, father and head of household). Data from 2009 to end of 2013 were included in this analysis. Nanoro has two main seasons: a rainy season from June to October and a dry season from November to May [24]. In this study, to reflect the malaria mortality seasonality and the potential lag effect of rainy season, the wet season was defined as July to November and the other seven months were defined as the dry season. There are 16 outpatient health facilities in the Nanoro health district and one inpatient health facility close to the village of Nanoro. There is also an inpatient health facility in Bousse just east of the district, which is the closest inpatient facility for some residents in the DSA, and therefore was included in this study (Figure 1).

**Figure 1:**
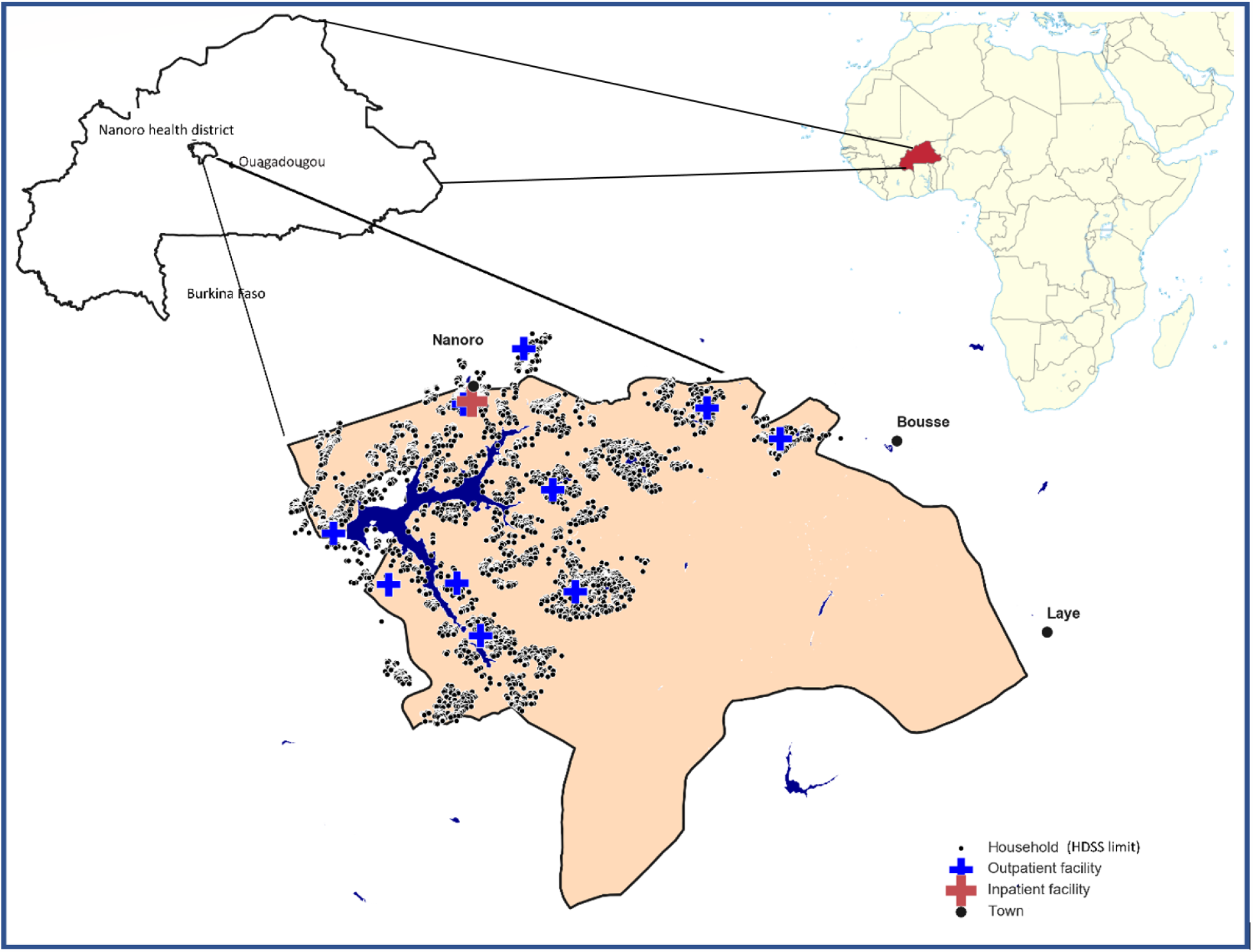
Nanoro health district is located in the rural center of Burkina Faso. Green dots represent the HDSS households and red crosses represent the health facilities.

Accessibility to both inpatient and outpatient health facilities was measured as Euclidean distance, travel time, and walking travel time. Travel time to the most accessible health facility was calculated using a global “friction surface” provided by the Malaria Atlas Project (MAP) at a resolution of 1 km for 2015, which estimates the travel time through every 1*×*1 km grid square on Earth using the fastest feasible surface travel [22]. A companion algorithm calculates the fastest journey time between any two user-provided points. This index may better capture the opportunity cost of travel than Euclidean or network distance, and reflects the information humans use to make transport decisions [22]. We also calculated walking travel time by modifying the friction surface developed by MAP, so that all roads received a fixed walking speed of 5 km per hour [22]. Fastest travel time was the main variable used to describe health-facility access in our models. Hereafter we will refer to this variable simply as “travel time.” Models using the other proximity variables are shown in Supplementary Material.

### Statistical Analysis

We estimated the survival probability of children under age five over the study’s nearly 5-year period, as 1 minus the product of average age-specific monthly survival rates from birth through 60 months, multiplied by 1000. Cox proportional hazards regression models [25] were used to estimate the association between under-5 survival and geographic, and seasonal risk factors. These include physical accessibility to health facilities, seasonality of death events during the survey, and age groups. The relationship between each of these factors and mortality risk was assessed one at a time as both categorical and continuous variables (when possible). The final multivariable model adjusted for risk factors that were significant on a univariate model and available for the entire dataset.

For each child, the follow-up time was taken as the time an individual was present within the age group during follow-up, which is the time from the date of first event in the survey, birth or enrollment or in-migration until age 5, out-migration, end of 2013, or death. Village was added as a cluster term to the model to estimate a robust variance. All the analyses and the mapping were performed in R [26].

## Results

### Demographics and Child Mortality

The key demographic characteristics of the study population are given in Table 1. At any given time during the study period, about 8,000 children under 5 years old lived in the district. Cumulatively 23,639 children were under age of 5 and in the district for at least part of the study period, contributing about 37,276 child-years of follow-up. Most children lived within 40 minutes of travel to an inpatient facility. Median household size was smaller among those living near an inpatient facility than those live further away (Table 1).

**Table 1.** Demographic characteristics of Nanoro HDSS for children under-5, 2009-2013

|  | Travel time to inpatient health facilities |  |  |  |
| --- | --- | --- | --- | --- |
|  | [0-20)min | [20-40)min | [40-60)min | 60+min |
| Number of deaths | 142 | 255 | 124 | 94 |
| Number of children | 7039 | 10266 | 3950 | 2596 |
| Household size | 6 (3,11)* | 7 (4,14) | 7 (4,13) | 9 (4,14) |
| Number of outpatient health facilities | 3 | 3 | 6 | 4 |
| Ratio of female to male children | 0.96 | 0.99 | 1.04 | 1.01 |
| Mother's age at birth in year | 27.1 (22.3,33) | 27.2 (22.4,32.7) | 27.2 (22.4,32.7) | 27.2 (22.4,32.5) |

\*Median and interquartile range.

The reported overall mortality rate among children under 5 in Nanoro HDSS during the study period was 64.9 deaths per 1,000 live births. Within the district, the village of Nanoro had the lowest under-5 mortality at 29.4 per 1,000 live births, approaching the 2030 SDG of *<*25. The southern and eastern edges of the DSA had substantially higher mortality rate (91.8 per 1,000 live births) (Supplementary Material, Figures S1-S3).

### Factors Associated with Child Mortality

The association between child mortality and the variables described in Methods is summarized in Table 2 and Figure 2. As expected, risk of death decreased with increasing age. Children between 1 to 2 years old were at lower risk of mortality by 48% (HR=0.52, 95% CI= 0.41-0.65) than infants, and the risk of mortality was lowest for children 3 to 4 years old (HR=0.39 vs. infants, 95% CI = 0.27-0.57) (Figure 2).

**Table 2.** Results of adjusted and unadjusted Cox regression models for under-5 mortality. Hazard Ratios are presented with 95% confidence intervals in parentheses.

| Parameters | Mortality per 1,000 live births | Unadjusted Hazard ratio | P-value | Adjusted Hazard ratio | P-value |
| --- | --- | --- | --- | --- | --- |
| **Age group (month)** |  |  |  |  |  |
| [0 – 12] | 327* | - | - | - | - |
| (12 – 24] | 136 | 0.50 (0.40, 0.63) | < 0.001 | 0.52 (0.41,0.65) | < 0.001 |
| (24 – 36] | 71 | 0.28 (0.20, 0.39) | < 0.001 | 0.29 (0.21,0.41) | < 0.001 |
| (36 – 48] | 56 | 0.39 (0.27, 0.56) | < 0.001 | 0.39 (0.27,0.57) | < 0.001 |
| (48 – 60] | 25 | 0.72 (0.48, 1.06) | 0.097 | 0.75 (0.49-1.13) | 0.17 |
| **Travel time to inpatient health facilities (min)** |  |  |  |  |  |
| [0 – 20) | 50.03 | - | - | - | - |
| [20 – 40) | 63.44 | 1.23 (0.89, 1.68) | 0.21 | 1.24 (0.89,1.71) | 0.20 |
| [40 – 60) | 79.49 | 1.51 (1.13, 2.03) | 0.006 | 1.52 (1.13,2.06) | 0.005 |
| 60+ | 88.51 | 1.74 (1.28, 2.38) | < 0.001 | 1.74 (1.27,2.40) | < 0.001 |
| **Seasonality** |  |  |  |  |  |
| Dry Season (Dec-June) | 64.49 | - | - | - | - |
| Wet Season (July-Nov) | 65.22 | 1.44 (1.15, 1.81) | 0.0018 | 1.28 (1.07, 1.53) | 0.0074 |
| **Travel time to outpatient health facilities (min)** |  |  |  |  |  |
| [0 - 5) | 58.11 | - | - | - | - |
| [5 – 10) | 66.99 | 1.15 (0.8, 1.65) | 0.44 |  |  |
| [10 – 15) | 69.72 | 1.20 (0.87,1.67) | 0.26 |  |  |
| [15 – 20) | 65.87 | 1.09 (0.75,1.59) | 0.64 |  |  |
| 20+ | 63.41 | 0.99 (0.58,1.69) | 0.99 |  |  |

\* Number of deaths within each age group.

**Figure 2:**
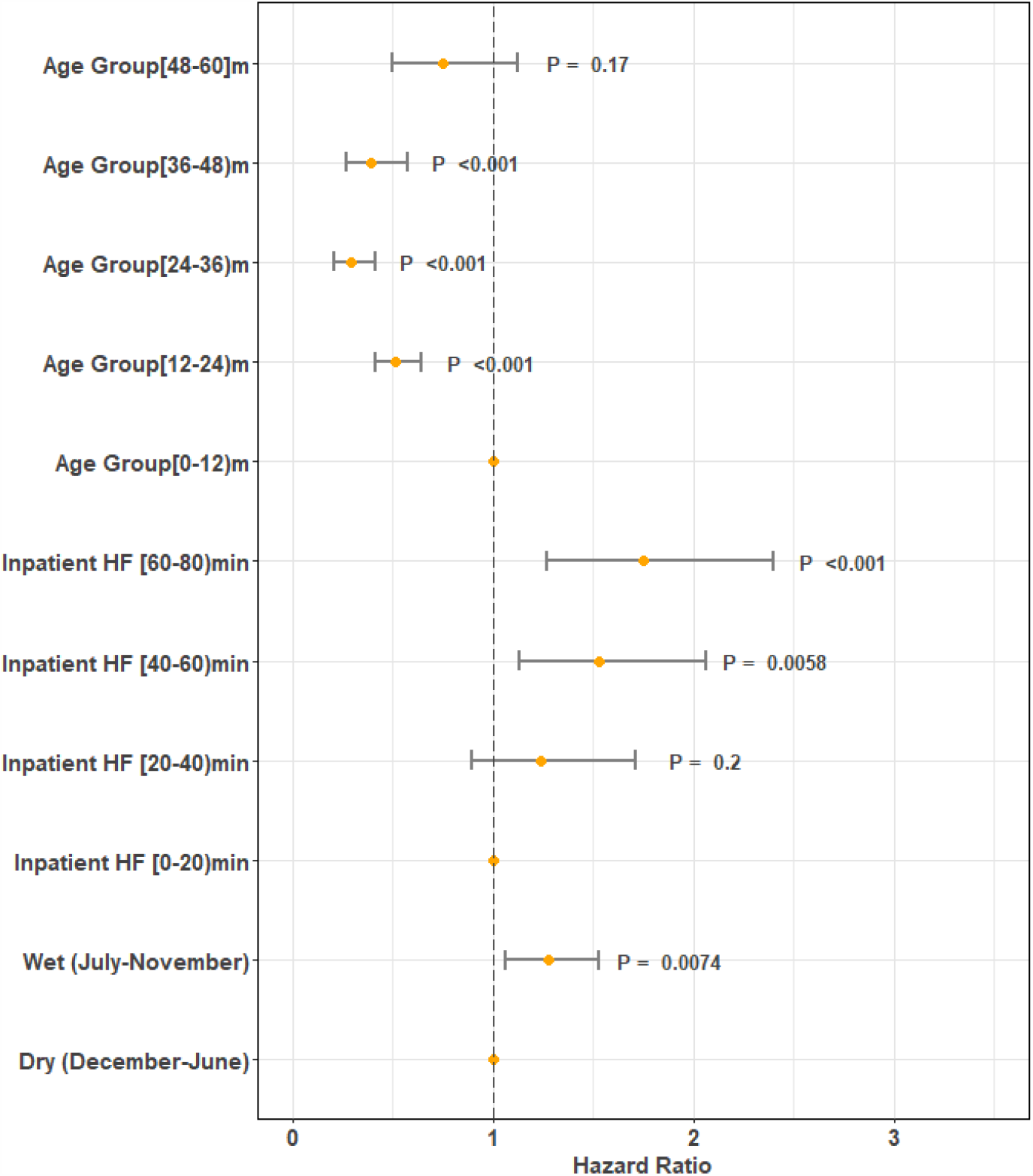
Hazard ratios of multivariable models associated with the probability of mortality of children under-5. Risk factors reducing the probability of death have hazard ratios lower than 1, to the left of the vertical dashed line. Hazard ratios (yellow points), 95% confidence intervals (horizontal lines) and p-values are shown. The variables with the yellow points on the vertical dashed line represent the reference groups.

### Seasonality

The under-5 mortality hazard was higher during the wet season (Jul-Nov) (HR=1.28, 95% CI = 1.07-1.53) than the dry season (Figure 2). Out-migration also had a clear seasonal pattern, and was higher during the dry season, highlighting the potential effect of out-migration on the child mortality pattern (supplementary material, Figure S8).

### Accessibility to Health Facilities

Under-5 mortality increased significantly with increasing travel time to an inpatient health facility (P<0.001). In particular, children living 40-60 minutes away from an inpatient facility experienced a 1.52 times higher mortality hazard (95% CI = 1.13-2.06) than those living within 20 minutes, and children living *>* 60 minutes away experienced a relative hazard of 1.74 (95% CI = 1.27-2.40) (Figure 2). Similar associations were found when using Euclidean distance or walking travel time (supplementary material, Figure S4 and Tables S1-S2). By contrast, there was no statistically significant association between accessibility to outpatient health facilities and under-5 mortality (supplementary material, Figure S5 and Tables S1-S2).

## Discussion

Our study provides insight into child mortality patterns in the Nanoro health district, Burkina Faso by linking it to various demographic, spatial and temporal risk factors. One distinction of our study is the evaluation of accessibility to both inpatient and outpatient health facilities. In the recent meta-analysis by Rojas-Gualdrand and Caicedo-Velazquez [18], the majority of studies included in its under-5 mortality endpoint estimate measured distance from any health center with no distinction between inpatient and outpatient. There were also inconsistencies regarding the effect of accessibility to health care on child and neonatal mortalities. In Malawi, DHS data showed no association between distance to delivery care and early neonatal mortality, and in Zambia, early neonatal survival was higher with increasing distance [27]. On the other hand, analysis of DHS data in Madagascar showed a higher risk of infant mortality among those who lived further from a health facility [28]. In rural western Burkina Faso, rural Ethiopia and Tanzania, accessibility to health facilities was found to be a major risk factor for infant, child and overall under-5 mortality [15, 16, 29]. Our analysis is in agreement with the latter studies, and indicates that impeded access to an inpatient health facility might be a major risk factor for child mortality. Our study also suggests that accessibility to outpatient health facilities does not drive the pattern of child mortality in the study area. We speculate that outpatient health facilities do not provide the level of care children need in a life or death situation. We note the confounding factor that inpatient health facilities are usually located in towns and major villages, with better food, water, and other living conditions for residents, as well as generally higher education and socioeconomic status. Another distinction of our study is the use of the recently developed global accessibility map that accounts for the spatial locations and properties of roads, railroads, rivers, water bodies, topographical characteristics, land cover, and national borders [22]. Accounting for these features leads to a more accurate measurement of accessibility than Euclidean or network distance that has been commonly used in previous studies.

There was a statistically significant association between seasonality of death and under-5 mortality, with the wet season having a higher mortality rate, reflecting the malaria mortality pattern.

Some of the limitations of our work are other risk factors that we have not accounted for and may be important to our outcomes, such as family wealth status, family health-seeking behavior, sanitation and hygiene information, and effects of flooding. Also, the travel time index we used in this study is based on the assumption that everyone could use the fastest travel method possible. However, our analysis using walking travel time showed a similar association with under-5 mortality. An additional limitation is that the friction surface developed by MAP does not account for the seasonal variation, which can affect travel time to health facilities. Last but not least, this is an observational study, and therefore any association is subject to potential confounding factors, as discussed above for inpatient facilities.

## Conclusions

Our study emphasizes that inequity in mortality rate is not only seen between rural and urban areas, but also within a relatively small rural area. It also highlights the importance of accessibility to health care in rural Burkina Faso. Our findings can help health policy makers and program developers in the health district and similar districts, to understand the potential effect of health infrastructure designs and the most effective locations of health facilities. Novel strategies, such as improved transportation to inpatient facilities during a child health emergency, strengthening of outpatient health facilities, and training community health workers in the rural area, are necessary for mitigating the physical limitations to accessing health care in the area. Also, reducing the socioeconomic inequalities between rural and urban areas as well as within each area, can help enhance access to health services for poor people and reduce child mortality [19, 34].

### Availability of data and materials

The datasets used and/or analyzed during the current study are available from the authors upon reasonable request. A public version of the dataset with fewer variables, and excluding household location, is available on the INDEPTH website.

## Abbreviations

SDG: Sustainable Development Goals
HDSS: Health and Demographic Surveillance System
CRVS: Civil Registration and Vital Statistics
DHS: Demographic and Health Surveys
INDEPTH: International Network for the Demographic Evaluation of Populations and Their Health in Developing Countries
CRUN: Clinical Research Unit of Nanoro
CMA: Centre Medicale avec Antenne Chirurgicale Saint Camille de Nanoro
DSA: Demographic Surveillance Area
MAP: Malaria Atlas Project

## Acknowledgements

The authors wish to thank the individuals of the Nanoro HDSS for their participation and their cooperation in this project, and the Ministry of Health of Burkina Faso for providing support.

## Funding

This work was supported by Bill and Melinda Gates through Global Good Fund.

## Author Information

Affiliations

^**1**^**Institute for Disease Modeling, Building IV, 3150 139th Ave SE, Bellevue, WA, USA, 98005**.

Navideh Noori, Assaf P. Oron, Edward Wenger, Andre Lin Ouedraogo

^**2**^**Institut de Recherche en Sciences de la Sante (IRSS)/Clinical Research Unit of Nanoro (CRUN), Nanoro, Burkina Faso**.

Karim Derra, Innocent Valea, Aminata Welgo, Toussaint Rouamba, Palwende Romuald Boua, Athanase SOME, Eli Rouamba, Hermann Sorgho, Halidou Tinto

^**3**^**Laboratory of Parasitology and Entomology, Centre Muraz, Bobo-Dioulasso, Burkina Faso**.

Innocent Valea, Halidou Tinto

## Contributions

NN, KD, IV, APO, ALO participated in the design of the analysis. NN and KD conducted the analysis, and NN drafted the original manuscript. All authors revised the manuscript. KD, IV, AW, TR, PRB, AB, ER, HS, HT designed the household survey and collected and cleaned the survey data. IV, APO, ALO supervised the project. HT, EW acquired funding and administered the project. The authors read and approved the final manuscript.

## Corresponding author

correspondence to.

## Ethics declarations

The ethical approval was waived by the Burkina Faso Ministry of Health, regarding the general population data of the HDSS (demographics and mortality data) used in this study.

## Consent of publication

Not applicable.

## Competing interests

The authors declare that they have no competing interests.

## Supplementary Information

Supplementary material.pdf

## Notes

### Competing Interest Statement

The authors have declared no competing interest.

